# Prevalence and Determinants of Musculoskeletal Symptoms Among Field Health Workers in Bin Qasim Town, Karachi

**DOI:** 10.64898/2026.05.03.26352346

**Authors:** Azqa Mazhar, Abdur Rasheed, Shayan Khakwani, Zahra Hoodbhoy

## Abstract

**Background:** Work-related musculoskeletal symptoms such as pain, stiffness, and swelling are a common occupational health issue that affect well-being and increase healthcare costs. Continuous physical effort, long hours of sitting, and poor awareness of proper ergonomics often lead to or worsen these conditions.

**Objective:** This study determined the frequency of musculoskeletal symptoms and the associated risk factors with musculoskeletal symptoms among field health workers in Bin Qasim Town, Karachi.

**Material & Methods:** A cross-sectional study was employed and collected data from Karachi based pre-urban communities i.e.: Ibrahim Hydri, Rehri Goth and Bhains Colony. Study duration was 9 months. MSK symptoms were assessed using the standardized Nordic Musculoskeletal Questionnaire (NMQ). Prevalence of MSK symptoms was assessed over 12 months and 7 days. Participants with pain in ≥2 regions of the upper or lower limbs were classified as having upper or lower limb symptoms, respectively. Multivariable logistic regression was used to identify associated factors for MSK symptoms in last 12 months.

**Results:** 132 participants were recruited. Most frequently reported pain region in the last 12 months was lower back 111(84%) and shoulder 81(61%). Similarly, the most affected region in the last 7 days was also lower back 39(29%) followed by shoulder 33(25%). Upper limb MSK symptoms were significantly associated with bachelor’s or higher educated (OR=3.38; 95% CI: 0.67–7.42), sitting 3–4 h/day (OR=3.46; 95% CI: 1.11–10.75), and walking 3–4 h/day (OR=2.88; 95% CI: 1.05–7.85). In lower limb, married workers had 2 times higher odds of lower limb MSK symptoms (OR=2.36; 95% CI:1.04 - 5.35), while those who worked > 30 hours/week had 67% lower odds of having lower limb MSK symptoms (OR=0.33, 95% CI:0.15 - 0.72).

**Conclusion:** Field health workers frequently reported MSK symptoms in both limbs. Preventive strategies such as ergonomic training, task rotation, and targeted support for married female workers are recommended to reduce the long-term impact.

## Introduction

Work-related musculoskeletal (MSK) symptoms have been identified as a newly developed occupational health issue due to their significant impact on human well-being and resultant healthcare expenditure (1). These symptoms include persistent discomfort, swelling, dull pain, and stiffness in different joints of the body to reduce functionality of life by making it challenging to do professional tasks and activities of daily living (2, 3). The symptoms can be caused by both professional factors (work-related characteristics, posture, ergonomics, psychosocial problems) and personal elements (age, gender, body mass index (BMI), physical inactivity, and tobacco use) (1).

Global estimates show that the prevalence of MSK conditions ranges between 20% and 33%, with the World Health Organization (WHO) estimating that 1.71 billion individuals worldwide suffer from Musculoskeletal Disorders (MSDs) (4). The WHO also identifies MSK conditions as one of the top causes of disability, ranking second in the Eastern Mediterranean Region (5). Literature suggests that MSDs accounts for 16% of the lifespan spent with disability and has a greater affinity for female gender and persons of working age (5). In developing nations, approximately one-fourth of people reported having MSK symptoms, which limit their ability to perform everyday tasks (6). In Asian countries, between 12% and 45% individuals have stated severe discomfort resulting from MSDs, with four times higher risk among healthcare providers compared to manufacturing workers (7). An Indonesian study reported that healthcare facility workforce regularly encountered ergonomics risks including muscular aches/sprains (76.5%), overstretching (67.5%), elbow/wrist/neck discomfort (56.0%), poor posture (56.0%), and bending/twisting (55.5%) (10). Furthermore, women account for 40% of the total workforce and are at a higher risk of experiencing MSK problems because they are subjected to more physical, functional, and psychological stress (6).

Field Health Workers (FHWs) are community level-staff who work for different projects to recruit patients, perform surveys, and conduct household visits to gather information, provide health services, or interventions in low-and middle-income countries (LMICs) such as Pakistan (8). FHWs face occupational challenges including atmospheric pollutants, exposure to humidity, sun radiation, infections, urban harassment, and work overload, which may lead to physical as well as mental health issues (9). Previous research has shown that these MSK symptoms are prevalent among community workers. For example, a Brazilian study found that 65.9% of female community workers had musculoskeletal symptoms in the last 12 months, with the lower back (65.9%) and neck (61.4%) being the most painful regions (10). In 2023, a systematic review concluded that workload is a significant barrier for community workers in LMICs (11), with 77.6% of studies reporting community workers feeling overburdened because of high workload. Limited transportation was reported in 25.6% of studies, frequently leading to long-distance walking and contributing to physical strain and musculoskeletal health issues (11). While musculoskeletal problems have been studied among doctors, nurses and other health care workers in LMICs, evidence among FHWs remains limited. Bridging this gap can identify the common body regions of musculoskeletal symptoms among FHWs, to improve their work ergonomics, and help reduce these symptoms.

Therefore, the purpose of this study is to explore the frequency of MSK symptoms among FHWs and to assess sociodemographic factors related to these symptoms. By examining the association between sociodemographic factors and MSK symptoms, we aim to enhance our understanding of the causes that contribute to MSDs in this population and propose targeted interventions.

## Materials And Methods

### Study design, Subjects and Population

An analytical cross-sectional study design was employed. Data was collected from 3 pre-urban communities of Bin Qasim Town of Karachi Pakistan i.e: Ibrahim Hydri (IH), Rehri Goth (RG) and Bhains Colony (BH) where field health workers are actively engaged in field settings, handling the management, oversight of site activities, operations and frequently visit patients’ homes to recruit them and collect data from them there. The duration of the study was 9 months (Nov 2024 – July 2025).

### Sample Size and Technique

The sample size calculation was performed using OpenEpi software version 3.0.1. By using anticipated frequency of MSDs as 46%(12), confidence interval as 95%, level of significance as 5%, and design effect as 1%, the calculated sample size was 132 FHWs. A purposive sampling technique was employed. The study team conducted interviews at health centers present in each field site in a private room, ensuring confidentiality that only the participant and the interviewer are present. The team approached the FHWs and educated them about the study protocol and MSK symptoms. They obtained written informed consent in Urdu; only those who provided the consent were considered as study participants.

### Eligibility Criteria

Field health workers aged 18 years and above, identified through official work Ids and working on field for more than 1 year and willing to participate in the study and give informed written consent. Participants with any physical disability, such as someone with polio, or any self-reported pre-existing degenerative diseases, such as osteoarthritis (OA) and rheumatoid arthritis (RA), were excluded from the study.

### Study Variables and Tool

A self-constructed questionnaire was designed to collect detailed information about the participants’ background characteristics including age, marital status, position title, education level and year of experience. Furthermore, information about the type of work, nature of work they perform, the amount of weight they carry at work, their monthly income, perception of workload and screen time exposure at work was also collected.

To identify MSK symptoms in FHWs the standardized Nordic musculoskeletal questionnaire (NMQ) was used. NMQ is an inexpensive tool for recognizing and evaluating musculoskeletal risk factors in workers. (13). It is a structured questionnaire, with binary and multiple-choice options and can be self-administered or utilized in interviews. It includes two sections: a demographic and work-related section, capturing age, work duration, height and weight, and a region-specific section focusing on questions related to the lower back and neck/shoulders. The goal of the general section is easy surveying, while the specific ones allow for detailed assessment, capturing information on symptom frequency, duration, severity, and functional impact (14).

**Section 1**: Is a questionnaire with 40 single best choice items to identify nine symptom sites: neck, shoulders, upper back, elbows, low back, wrist/hands, hips/thighs, knees, and ankles/feet. Respondents are asked if they have experienced any musculoskeletal issues in the past 12 months and 7 days that have restricted normal physical activity.

**Section 2**: Includes additional questions about the neck, shoulders, and lower back, covering important topics. Twenty-five forced-choice questions investigate accidents, functional impact at work and home, the duration of the problem, health professional assessment, and musculoskeletal difficulties during the last week. (15)

The NMQ has been widely used to access musculoskeletal symptoms and discomfort in various groups, including healthcare workers (18–26) In Pakistan, it has been applied among healthcare workers, including dental practitioners to evaluate work related MSK symptoms, with findings indicating a high prevalence of neck and lower back pain and notable impacts on occupational functioning (16). Reliability studies report internal consistency exceeding 70% (13), while validity assessments comparing NMQ responses with clinical evaluation demonstrate 80-100% agreement. The questionnaire has also shown good diagnostic performance, with sensitivity ranging from 66-92% and specificity from 71-88% for identifying self-reported MSK symptoms (15).

### Statistical analysis

Stata version (17.0) and R Studio software (4.2.3) were used for statistical analysis. Categorical variables were reported as frequency and percentages. Prevalence of MSK pain in 9 body regions was reported in the last 12 months and 7 days. Binary logistic regression analysis was applied separately on upper limb and lower limb to assess the association of MSK symptoms and potential associated factors. Females experiencing musculoskeletal (MSK) pain in ≥ 2 upper limb regions were classified as having upper limb symptoms: similarly, those with pain in ≥ 2 lower limb regions. The associated risk factors with P-value ≤ 0.25 in univariable analysis were eligible for the multivariable model, where p-value ≤ 0.05 was considered statistically significant. Both crude and adjusted odds ratios (OR) along with 95% confidence intervals (CI) were reported.

### Ethical consideration

The approval from Aga Khan University’s Ethics Review Committees (ERC#2024-10524-31273) was also obtained. All participants provided informed written consent, and necessary measures were taken to ensure the confidentiality and privacy of the gathered information.

## Results

We included 132 FHWs in this study. Most of the participants (n=67, 50.8%) were aged 30-39 years (n=67, 50.7%), which was followed by 20-29 years (n=43, 32.5%) and 40 & above (n=22, 16.6%). Most of them were married (n=77, 58.3%) and had field experience of more than 5 years (n=70, 53.0%). More than two thirds (n=89, 67.4%) had an income of greater than 40,000 PKR per month, with less than half (n=55, 41.6%) having > 30 average working hours per week. The demographic characteristics of the participants are shown in Table 1

**Table:1.** Sociodemographic characteristics of Female Health Workers.

| **Characteristic** | **N = 132** |
| --- | --- |
|  | **n (%)** |
| **Age** |  |
| 20-29 | 43 (32.6%) |
| 30-39 | 67 (50.8%) |
| 40+ | 22 (16.7%) |
| **Marital Status** |  |
| Married | 77 (58.3%) |
| Unmarried | 55 (41.7%) |
| **Education** |  |
| Primary | 28 (21.2%) |
| Secondary | 46 (34.9%) |
| Bachelor's & above | 58 (43.9%) |
| **Years of Field Experience** |  |
| 1-2 years | 25 (18.9%) |
| 3-5 years | 37 (28.0%) |
| More than 5 years | 70 (53.0%) |
| **Type of work** |  |
| Household Visits only | 105 (79.6%) |
| Clinic Site & Household both | 27 (20.5%) |
| **Sitting Hours** |  |
| 1-2 hours | 95 (72.0%) |
| 3-4 hours | 37 (28.0%) |
| **Standing Hours** |  |
| 1-2 hours | 78 (59.1%) |
| 3-4 hours | 0 (0.0%) |
| Not standing | 54 (40.91%) |
| **Walking Hours** |  |
| 1-2 hours | 54 (40.9%) |
| 3-4 hours | 78 (59.1%) |
| **Weight Carry** |  |
| < 1 kg | 39 (29.6%) |
| 1-5 kg | 67 (50.8%) |
| >6 kg | 26 (19.7%) |
| **Perception of Workload** |  |
| Low to Moderate | 88 (66.7%) |
| High | 44 (33.3%) |
| **Screen Time hours/day** |  |
| ≤ 2 hours | 44 (33.3%) |
| > 2 hours | 88 (66.7%) |
| **Experience of Current job** |  |
| ≤3 Years | 70 (53.0%) |
| >3 Years | 62 (47.0%) |

| **Working hours/week** |  |
| --- | --- |
| ≤30 hours per week | 77 (58.3%) |
| >30 hours per week | 55 (41.7%) |
| **Dominant Side** |  |
| Right-Handed | 124 (93.9%) |
| Left-Handed | 8 (6.1%) |
| **BMI** |  |
| Underweight (< 18.5) | 29 (22.0%) |
| Normal (18.5-24.9) | 45 (34.1%) |
| Overweight (25-29.9) | 33 (25.0%) |
| Obese (≥30) | 25 (18.9%) |

The prevalence of MSK symptoms varied across nine anatomical regions during the last 12 months and last 7 days. The most frequently reported region in the last 12 months was lower back (n=111, 84.1%), subsequently followed by shoulder (n=81, 61.4%). The least reported region was the upper back (n=6, 4.6%). Similarly, the most affected region in the last 7 days was also lower back (n=39, 29.6%) followed by shoulder (n=33, 25.0%) with no reported symptoms in the elbow region. A diagrammatic illustration representing the prevalence of MSK symptoms in different body regions within the last 12 months and 7 days shown in (Fig 1-Fig 2).

**Figure:1.**
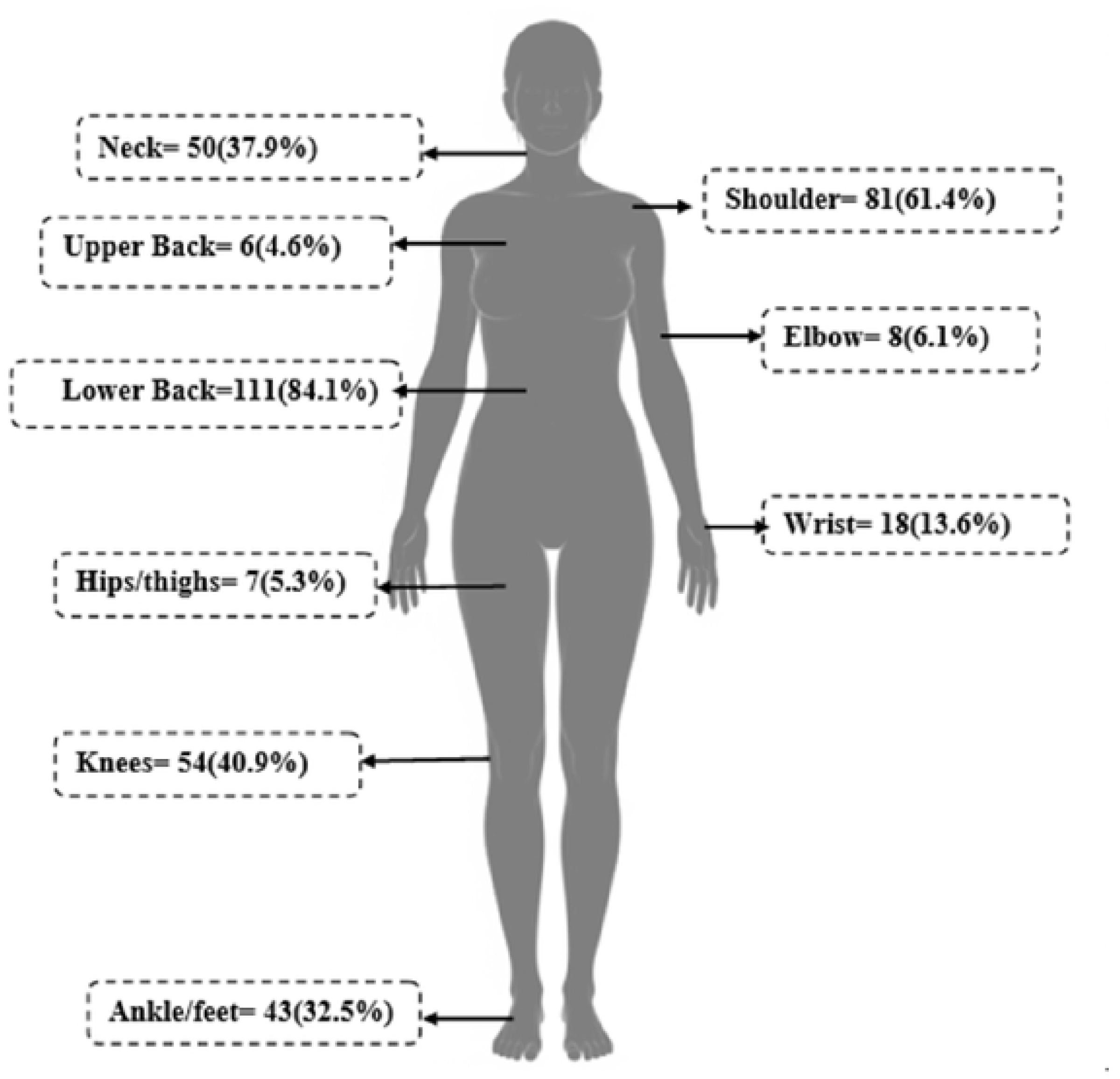
Distribution of MSK symptoms in last 12 months.

**Figure:2.**
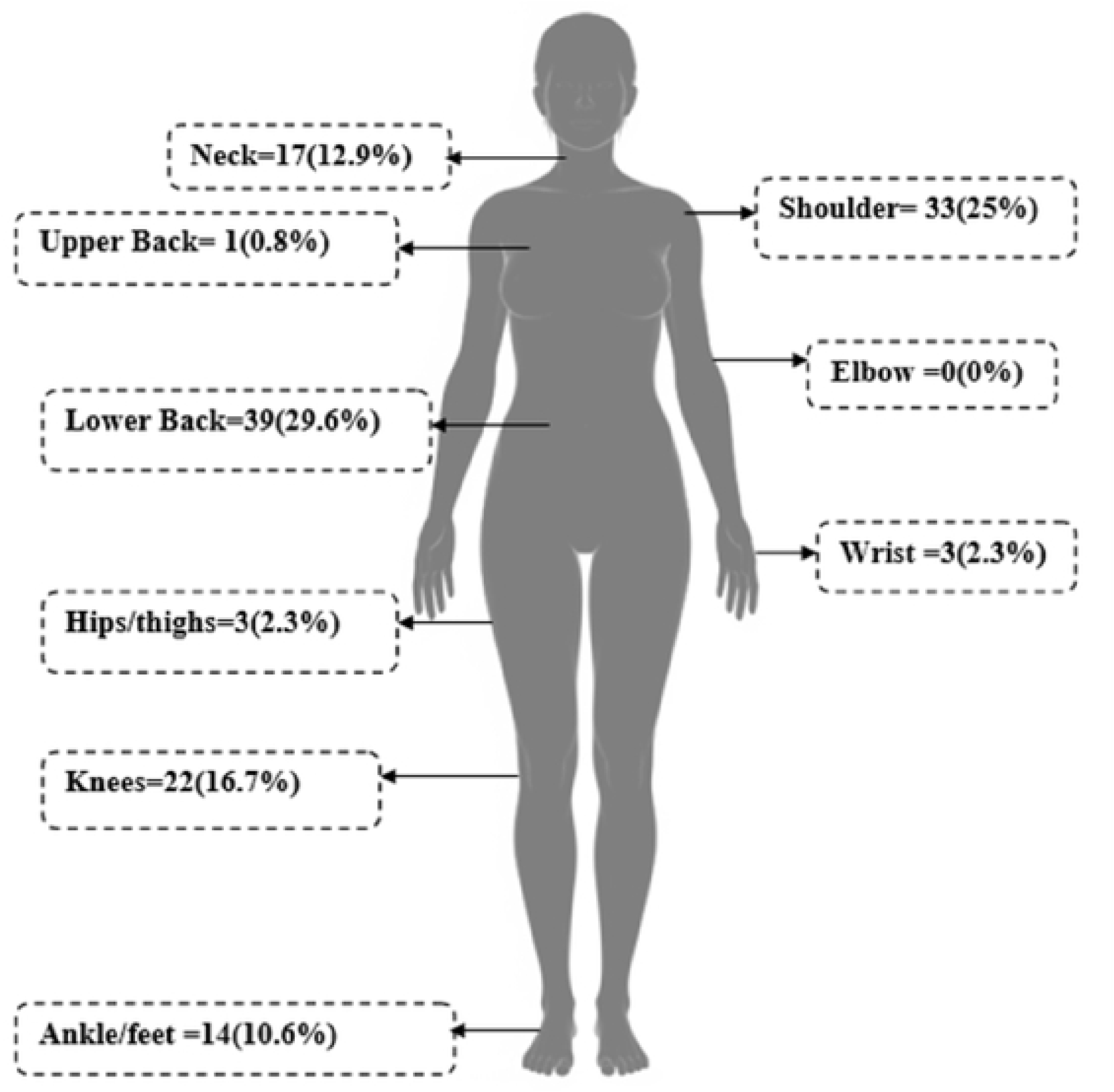
Distrubution of MSK symptoms in last 7 days.

Those who had upper limb pain, had a significant association with education (OR:3.38; 95% CI: 1.02-11.15) compared to those who were less educated, sitting 3-4 hours/day (OR: 3.46; 95% CI: 1.11 - 10.75), walking 3-4 hours/day (OR: 2.88; 95% CI:1.05 - 7.85).A marginally significant relationship was observed with BMI both among those who are underweight (OR: 2.63;95% CI:0.81 - 8.49) and overweight (OR: 2.83; 95% CI:0.90 - 8.93) (Table 2)

**Table:2.** Multivariable Logistic Regression Analysis.

| Regions | Upper Limb |  | Lower Limb |  |
| --- | --- | --- | --- | --- |
| Variable | Crude OR<br>(95% CI) | Adjusted OR<br>(95% CI) | Crude OR<br>(95% CI) | Adjusted OR<br>(95% CI) |
| **Age** |  |  |  |  |
| **20-29** | 1 | 1 | 1 | 1 |
| **30-39** | 2.84 (1.20 - 6.68) * | 1.42 (0.50 - 4.04) | 1.76 (0.80 - 3.82) | 1.15 (0.48 - 2.57) |
| **40+** | 3.3 (1.10- 9.86) * | 1.59 (0.38 - 6.64) | 0.87 (0.31 - 2.44) | 0.46 (0.13 - 1.60) |
| **Marital Status** |  |  |  |  |
| **Unmarried** | 1 | - | 1 | 1 |
| **Married** | 1.42 (0.69 -2.90) | - | 1.98 (0.98 - 4.01) | 2.36 (1.04 - 5.35)<br>* |
| **Education** |  |  |  |  |
| **Primary** | 1 | 1 | 1 | - |
| **Secondary** | 1.14 (0.40 - 3.22) | 2.23 (0.67 - 7.42) | 1.01 (0.39 - 2.62) | - |
| **Bachelor's & above** | 2.29 (0.86 - 6.06) | 3.38 (1.02- 11.15) * | 1.35 (0.54 - 3.38) | - |
| **Years of Field Experience** |  |  |  |  |
| **1-2 years** | 1 | 1 | 1 | - |
| **3-5 years** | 0.78 (0.25 - 2.38) | 0.73 (0.20 - 2.66) | 0.59 (0.20 - 1.68) | - |
| **More than 5 years** | 2 (0.76 -5.25) | 1.6 (0.52 - 4.92) | 0.66 (0.26 - 1.71) | - |
| **Type of work** |  |  |  |  |
| **Clinic Site & Household both** | 1 | - | 1 | - |
| **Household Visits only** | 1.13 (0.47 2.71) | - | 1.19 (0.51 - 2.77) | -<br>- |
| **Sitting Hours** |  |  |  |  |
| **1-2 hours** | 1 | 1 | 1 | 1 |
| **3-4 hours** | 3.73 (1.68 - 8.27) * | 3.46 (1.11 - 10.75) * | 1.73 (0.78 - 3.80) | 1.85 (0.73 - 4.65) |
| **Standing Hours** |  |  |  |  |
| **No standing** | 1 | - | 1 | - |
| **1-2 hours** | 0.9 (0.44 - 1.84) | - | 1.43 (0.71 - 2.89) | - |
| **Walking Hours** |  |  |  |  |
| **1-2 hours** | 1 | 1 | 1 | - |
| **3-4 hours** | 0.54 (0.75 - 3.17) | 2.88(1.05 - 7.85) * | 0.98 (0.48 - 1.97) | - |
| **Weight Carry** |  |  |  |  |
| **< 1 kg** | 1 | - | 1 | - |
| **1-5 kg** | 0.67 (0.30 - 1.51) | - | 1.76 (0.34 - 1.69) | - |
| **>6 kg** | 1.01 (0.40 - 3.00) | - | 0.94 (0.34 - 2.59) | -<br>- |
| **Perception of Workload** |  |  |  |  |
| **Low to Moderate** | 1 | - | 1 | - |
| **High** | 1.09 (0.52 - 2.30) | - | 0.83 (0.40 - 1.72) | - |
| **Screen Time hours/day** |  |  |  |  |
| **≤2 hours/day** | 1 | 1 | 1 | - |
| **>2 hours/day** | 1.62 (0.76 - 3.48) | 1.69 (0.67 - 4.26) | 0.68 (0.32 - 1.44) | - |
| **Current job Experience** |  |  |  |  |
| **≤3 years** | 1 | - | 1 | 1 |
| **>3 years** | 1.22 (0.60 - 2.46) | - | 0.58 (0.29- 1.17) | 0.53 (0.25 - 1.13) |
| **Working hours/week** |  |  |  |  |
| **≤30 hours per week** | 1 | - | 1 | 1 |
| **>30 hours per week** | 1.18 (0.58 - 2.41) | - | 0.388 (0.19 - 0.79) | 0.33 (0.15 - 0.72) |
| **BMI** |  |  |  |  |
| **Normal (18.5-24.9)** | 1 | 1 | 1 | - |
| **Underweight (<18.5)** | 2.8 (0.99 -7.98) | 2.63 (0.81 - 8.49) | 0.81 (0.320-2.07) | - |
| **Overweight (25-29.9)** | 4.25 (1.56 - 11.5) * | 2.83 (0.90 - 8.93) | 1.18 (0.48 - 2.93) | - |
| **Obese (≥30)** | 5.09 (1.56 - 14.9) * | 2.83 (0.79 - 10.18) | 1.550.56 - 4.24) | - |
| *Indicates statistically significant variables (p-values < 0.05) |  |  |  |  |

In lower limb multivariable model, a significant association was found with marital status, which had 2 times increasing odds of lower limb pain in those who were married (OR: 2.36; 95%CI:1.04 - 5.35) compared to unmarried women. A protective relation was also shown among those who worked more hours, as odds of lower limb pain decreased among those who worked > 30 hours/week (OR: 0.33, 95%CI:0.15 - 0.72) compared to those who worked ≤30 hours/week. Maybe those who worked for a longer duration might hold supervisory roles that are less physically demanding. (Table 2)

Additionally, details on functional Impact and Healthcare Utilization for MSK symptoms were compiled from Section 2 of the NMQ, specifically focused on neck, shoulder, and lower back regions (Table-3)

**Table 3:** Functional Impact and Healthcare Utilization Due to Neck, Shoulder, and Lower.

| Impact Variable | Neck n (%) | Shoulder n (%) | Low Back n (%) |
| --- | --- | --- | --- |
| Ever had pain | 61 (46.2%) | 89 (67.4%) | 118 (89.3%) |
| Injured<br>(Hospitalized/Accident) | 5 (3.8%) | 9 (6.8%) | 4 (3.0%) |
| Job change | 0(0%) | 0(0%) | 8 (6.1%) |
| Pain duration of 12 months |  |  |  |
| No pain | 11 (8.3%) | 8(6.1%) | 7(5.3%) |
| 1-7 days | 23(17.4%) | 22(16.7%) | 28 (21.2) |
| 8-30 days | 2(1.5%) | 5(3.8%) | 13 (9.8%) |
| +30 days (not every day) | 24(18.2%) | 44 (33.3%) | 56 (42.2%) |
| Everyday | 1(0.8%) | 10 (7.6%) | 14 (10.6%) |
| Activity Reduced in 12 months |  |  |  |
| Work | 29 (22.0%) | 39 (29.5%) | 47 (35.6%) |
| Leisure | 21(15.9%) | 42 (31.8%) | 64 (48.5%) |
| Seek Care (doctor, physiotherapist, chiropractor etc.) | 9 (6.8%) | 10 (7.6%) | 37 (28.0%) |

Out of 132 female participants, lower back was the most reported site of pain (n=118, 89.3%), followed by shoulder (n=89, 67.4%) and neck (n=61, 46.2%). Specifically, injuries that required hospitalization were also reported, with highest report for shoulder region (n=9, 6.8%). Consequentially, only a small number of females (n=8, 6.1%) desired to change their jobs owing to lower back discomfort.

Lower back pain was the highest reported pain in the last 12 months, with almost half (n= 56, 42.2%) of participant experiencing lower back discomfort for more than 30 days. Lower back pain was also the primary cause of reduced functionality, with more than one third (n=47, 35.7%)_of participants reporting limited in work-related tasks and approximately half (n=64, 48.5%) reporting decreased participation in leisure activities due to pain. However, only about one-third (n=37, 28.0%) of participants reported seeking medical care for lower back pain.

## DISCUSSION

The purpose of this study was to explore the frequency of MSK symptoms among FHWs and to assess risk factors related to these symptoms. Overall, 39.3% had upper limbs and 55.3% had lower limb MSK symptoms. Factors such as high education level, marital status, prolonged sitting, walking and working hours were associated with MSK symptoms of upper and lower limb.

Our study identified that the most reported pain region in the last 12 months as well in last 7 days was lower back pain, which is consistent with prior literature. One European study comparing 188 healthcare workers and 151 administrative workers reported that lower back pain was the most frequent region of complaint and was also a significant reason of change in job position (17). The high burden of lower back pain observed across both low- and high-income countries may be largely attributed to the inherent physical demands of patient care, which frequently compromises optimal ergonomic positioning (18).Healthcare workers are routinely required to perform tasks such as equipment lifting and repositioning, often in constrained clinical environments that limit adherence to recommended ergonomic practices (18). In LMICs, additional factors such as long travel distances for home visits, challenging terrain, and exposure to poverty and violence may further exacerbate cumulative strain among healthcare workers (19, 20). These factors highlight the need for targeted strategies to reduce lower back strain among healthcare workers.

Education seemed to play an important role with FHWS reporting higher symptoms. This result is consistent with a middle eastern study conducted on 310 physiotherapists, which reported that cervical is most common pain site and those with a 5–6-year bachelor’s degree or postgraduate education had high work-related musculoskeletal disorders (21). Another Egyptians study on 401 physical therapists showed that higher education and working in multiple facilities is associated with upper back injuries and neck dysfunction (22). In contrast, a cross-sectional study of 334 registered female nurses in Malaysia found that work-related musculoskeletal disorders in neck, shoulder, and upper-limb pain, were highly prevalent and lower education level like diploma was significantly associated with high prevalence of MSD (23). These differences imply that the association between education and MSK symptoms is impacted by workplace ergonomics and requirements among occupations.

Our study also found a relationship between prolonged sitting and walking with upper limb symptoms, which is supported by previous studies indicating that prolonged and static sitting with incorrect posture puts strain on muscles of upper back and reduces blood flow that leads to pain in upper limb regions (24–26). While walking is normally considered beneficial for musculoskeletal health, a Japanese study found that walking while carrying items increased the risk of MSK symptoms in neck, shoulders, and elbows (27). This indicates that occupational walking, particularly associated with carrying heavy objects or repeated duties, which is routinely performed by these FHWs as they carry heavy equipment such as anthropometric instruments along with other materials to households, hence predisposing them to increased upper-limb strain.

In our study, married females had a significant relationship with lower-limb musculoskeletal problems. This finding aligns with a previous study which revealed that married females are more likely to have work related MSK pain (28). This increased risk may be related to the ‘twice the load’ of professional duties and considerable household and children responsibilities (5). This increases the physical and psychological burden on FHWs which may lead to MSK conditions.

In contrast, the significant inverse odds between prolonged working hours and lower limb MSK symptoms represents a unique finding that requires additional investigation, as it contradicts the current occupational health trends where longer working hours frequently increase MSK pain (29)

### Strengths And Limitations

This study has several strengths and limitations. The utilization of the validated standardized NMQ in a community-based setting improves its external validity. Additionally, the study is unique in concentrating on FHWs, who are an important part of the healthcare system, but may be neglected in health care worker assessments. Although the sample size was adequate for assessing frequency of MSK, it was not powered to evaluate associated factors. The cross-sectional design further limits causal interpretation. Some inverse relationships and large confidence intervals should be interpreted with caution.

### Conclusion

FHWs frequently report MSK symptoms in both upper and lower limbs. Preventive interventions are important for reducing the long-term impacts and risk factors associated with MSK symptoms among field workers. Additionally targeted support for married female health workers who frequently balance job tasks with significant household duties might help reduce dual workload stress. In combination, these strategies can help to reduce the likelihood, incidence, and development of MSK diseases in the future.

## Data Availability

data will be made available upon request

## Acknowledgments

The authors are appreciative to the participants and workers for their assistance and significant contributions to this study.

## Notes

### Competing Interest Statement

The authors have declared no competing interest.

### Funding Statement

The authors recieved no specific funding for this work

### Author Declarations

Ethical approval was obtained from the Ethical Review Committee of Aga Khan University, Karachi, Pakistan (Reference No: 2024-10524-31273). The study was approved on 10 October 2024 for a period of one year. The committee reviewed the protocol and determined that there were no major ethical concerns. All required study documents, including consent forms and questionnaires, were approved by the ERC.

